# Comparative Effectiveness of Alternative Intervals between First and Second Doses of the mRNA COVID-19 Vaccines: a Trial Emulation Approach

**DOI:** 10.1101/2023.03.06.23286848

**Authors:** Kayoko Shioda, Alexander Breskin, Pravara Harati, Allison Chamberlain, Benjamin A Lopman, Elizabeth T. Rogawski McQuade

## Abstract

**Importance:** mRNA COVID-19 vaccines require two primary doses. The optimal timing of second dose administration with respect to vaccine effectiveness of the primary series has not been thoroughly evaluated and has implications for vaccination strategies.

**Objective:** To assess whether the effectiveness of mRNA COVID-19 vaccines (Pfizer-BioNTech and Moderna) against SARS-CoV-2 infection differs by varying intervals between the first and second doses of the primary series among the general population.

**Design:** We employed a trial emulation approach (clone-censor weight analysis) to estimate the risk of SARS-CoV-2 infection after the first dose administration under the scenario where the total study population had followed each of the following interdose intervals: recommended by the Food and Drug Administration (FDA) (17-25 days for Pfizer-BioNTech; 24-32 days for Moderna), late-but-allowable (26-42 days for Pfizer-BioNTech; 33-49 days for Moderna), and late (≥43 days for Pfizer-BioNTech; ≥50 days for Moderna).

**Setting:** Georgia, USA.

**Participants:** Individuals who received ≥1 dose of mRNA COVID-19 vaccines in Georgia between December 13, 2020 and March 16, 2022.

**Exposure:** Dosing protocols based on the timing of the second dose administration.

**Main Outcomes and Measures:** SARS-CoV-2 infection was defined as a positive result by a real-time reverse transcriptase PCR or antigen test. The follow-up period began the day after the first dose administration and ended at the earliest of SARS-CoV-2 infection, protocol nonadherence, or end of study.

**Results:** In the short-term, the cumulative risk of SARS-CoV-2 infection was lowest under the FDA-recommended protocol (risk ratio (RR) on Day 50 after the first dose administration compared to the FDA-recommended protocol: 1.08 [95% confidence interval 1.07-1.10] under the late-but-allowable and 1.14 [1.12-1.16] under the late protocol). Longer-term, the late-but-allowable protocol resulted in the lowest risk (RR on Day 120: 0.83 [0.82-0.84] for the late-but-allowable and 1.10 [1.08-1.12] for the late protocol). The late protocol consistently yielded the highest risk among all protocols.

**Conclusions and Relevance:** Delaying the timing of the second dose administration by a week may provide stronger protection against SARS-CoV-2 infection, but a longer delay would increase the risk of infection.

**KEY POINTS:** *Question:* Does the effectiveness of mRNA COVID-19 vaccines differ by intervals between the first and second doses of the primary series?

*Findings:* This study of >6 million mRNA COVID-19 vaccine recipients in Georgia, US used a trial emulation approach to compare the risk of SARS-CoV-2 infection under three protocols based on the timing of the second dose (“recommended,” “late-but-allowable,” and “late”). The late-but-allowable protocol led to the lowest cumulative risk in a long term.

*Meaning:* Delaying the receipt of the second dose by a week may decrease the risk of SARS-CoV-2 infection, but a longer delay would increase the risk.

## INTRODUCTION

Two mRNA COVID-19 vaccines (Pfizer-BioNTech and Moderna) are currently authorized and fully approved for the prevention of COVID-19 in the United States.^1,2^ The interval between the 2-doses of the primary series recommended by the Food and Drug Administration (FDA) is three weeks for Pfizer-BioNTech and four weeks for Moderna.^3,4^ While the majority of mRNA vaccine recipients in the U.S. received their primary doses close to these recommended timings, some missed the second dose or received it outside the recommended interval.^5^ There were substantial differences in the completion of the primary series and adherence to the recommended schedule by race, ethnicity, age, and/or jurisdiction.^5^

Vaccine dosing schedules have an important public health relevance, especially in resource-limited settings. When facing a shortage of vaccine supply while experiencing high level of SARS-CoV-2 transmission, countries have considered delaying the administration of the second dose of mRNA vaccines as a pragmatic approach to achieve a higher coverage of the single dose in the population.^6^ For example, the U.K. delayed the timing of second dose administration from three weeks to twelve weeks.^7^ Because of the high relevance to public health, not only the immunogenicity^8^ but also the total public health impact of different dosing protocols should be rigorously estimated to inform policy decisions.

The variability of vaccine effectiveness or efficacy against SARS-CoV-2 infection by different timing of second dose administration was evaluated. Test-negative design (TND) studies in Canada found that, compared to the manufacturer-specified 3-4-week interval between the first and second doses, a 7-8-week interval increased the effectiveness against SARS-CoV-2 infection among the general population in British Columbia and Quebec^9^ and healthcare workers in British Columbia.^10^ In contrast, no evidence of different effectiveness by dosing interval was found in longitudinal studies among households^11^ and healthcare workers in the U.K.^12^ While these findings provide important insights, the underlying study designs have limitations, such as the potential for selection bias for TND studies.^13^ Also, previous studies compared effectiveness based on the incidence of COVID-19 only after people received the second dose. This could be misleading because longer interdose intervals result in longer time at the lower level of protection afforded by a single dose, increasing the risk of infection during the interdose interval. In order to identify the optimal dosing protocol, it is critical to compare the risk of infection not only after people are fully vaccinated but also throughout the whole course of vaccination including the interdose interval.

We used a trial emulation approach to evaluate the effectiveness of dosing protocols based on different interdose intervals of the mRNA COVID-19 vaccines. We used state-wide COVID-19 case and vaccination data in Georgia, US from 2020-2022. A trial emulation approach accounted for the duration of time at the sub-optimal levels of protection experienced during the inter-dose interval, while avoiding common biases that can occur when the date of treatment receipt and the start of follow-up differ.^14^ We also used the Cox proportional hazards (PH) model to compare the hazard of SARS-CoV-2 infection after the completion of primary doses.

## METHODS

### Study population

Our study population included individuals who received at least one dose of an mRNA COVID-19 vaccine between December 13, 2020 and March 16, 2022 in Georgia, US. Unvaccinated people were excluded because our goal was to evaluate vaccine effectiveness depending on different interdose intervals. See eMethods 1 for more information on excluded individuals.

### Data source

We extracted the information on the vaccine manufacturer, the date of receipt of each vaccine dose, demographic characteristics (age, gender, race, and ethnicity), and geographic region of residency (18 public health districts of residency^15^) from the Georgia Department of Public Health (GDPH) vaccine database. We also extracted SARS-CoV-2 test results for our study population from the State Electronic Notifiable Disease Surveillance System (SendSS), an electronic database to track patients with notifiable diseases, including COVID-19 cases, across Georgia. Data are reported to the GDPH from laboratories, hospitals, and providers through SendSS and/or Electronic Laboratory Reports (ELR).

### Exposure and outcome

The protocols under investigation in our analysis were defined based on the timing of the second dose administration of an mRNA COVID-19 vaccine relative to the first dose. We used the following four categories to characterize different interdose intervals: shorter than the FDA-recommended interval (≤16 days for Pfizer-BioNTech and ≤23 days for Moderna; “early” protocol), the FDA-recommended interval (17-25 days for Pfizer-BioNTech and 24-32 days for Moderna; “recommended” protocol), longer than the FDA-recommended interval but within the allowable interval (26-42 days for Pfizer-BioNTech and 33-49 days for Moderna; “late-but-allowable” protocol), and after the FDA-recommended interval and the allowable interval (≥43 days for Pfizer-BioNTech and ≥50 days for Moderna; “late” protocol).^5^

Our outcome was SARS-CoV-2 infection defined as a positive result of real-time reverse transcriptase PCR test or antigen test. For the trial emulation approach, the follow-up period began the day after the index date (i.e., the day each individual received their first dose of mRNA COVID-19 vaccine) and ended at the earliest of SARS-CoV-2 infection, protocol nonadherence, or end of study (March 16, 2022). For the Cox PH model, the index date was the date of the second dose administration and ended at the earliest of SARS-CoV-2 infection or end of study (March 16, 2022).

### Covariates

We included demographic characteristics (age, sex, race, and ethnicity), public health districts of residence, vaccine manufacturers, and the presence of reported COVID-19 infection before vaccination to account for confounding in the analysis. We also adjusted for the calendar month and year of the first dose of vaccination to account for changing levels of community transmission throughout the pandemic, varying state- and local-level SARS-CoV-2 prevention policies over time (e.g., mask mandates), and the different severity and transmissibility of SARS-CoV-2 variants.

### Trial emulation: clone-censor weight analysis

We employed a trial emulation approach (clone-censor-weight analysis) to understand how the different intervals between the first and second doses of the primary series of mRNA COVID-19 vaccines may change the risk of SARS-CoV-2 infection after the first dose administration.^14,16,17^ This method mimics a per-protocol analysis of a randomized controlled trial in which individuals are randomly allocated to alternative dosing protocols.

Adopting the methods used by Butler, *et al*.,^18^ we created three copies of the longitudinal dataset corresponding to the three mRNA COVID-19 vaccination protocols of interest (FDA-recommended, late but allowable, and late). This method addresses measured confounding at baseline because the copies of each observation are identical at the start of follow-up. In each protocol-specific copy, a vaccine recipient who did not follow a given protocol was considered nonadherent and was censored at the time their vaccination course differed from the protocol (eMethods 1, eFigure 2, eTable 1). For example, a patient who received their second dose of the Moderna vaccine 30 days after their first dose would be censored on day 30 for the ‘late-but-allowable’ protocol. The third dose (booster dose) administration was considered nonadherence to the protocol and individuals were censored on the day of the booster dose administration. Informative censoring due to protocol non-adherence was addressed with inverse probability of censoring weights (IPCW) that account for the aforementioned covariates. The weights were designed to upweight individuals who remain adherent to the vaccine protocol at each time to have the same covariate distribution as the entire study population, thus creating a weighted population that represents the entire study population had all individuals remained adherent to the certain vaccine protocol throughout follow-up.

For each of the three protocol-specific copies, we calculated the cumulative risk functions of SARS-CoV-2 infection had the total study population followed the corresponding protocol. We calculated the risk ratio (RR), setting the FDA-recommended protocol as a reference. We computed 95% confidence intervals (CIs) using a nonparametric bootstrap based on 200 resamples.^19,20^

### Cox PH model analysis

To compare to a more conventional analysis, we fit Cox PH models to compare the hazard of SARS-CoV-2 infection after the administration of the *second* dose of mRNA COVID-19 vaccines between interdose interval variations. In contrast to the trial emulation, the survival time for this analysis was the number of days between the date of the *second* dose receipt and the date of SARS-CoV-2 infection or censoring. We estimated adjusted hazard ratios (aHRs) controlled for the aforementioned covariates, setting the “recommended” interval as a reference. The model was stratified by the presence of SARS-CoV-2 infection prior to the vaccination, because Kaplan-Meier survival curves for individuals with and without prior infection crossed around 275 days, invalidating the proportional hazards assumption for that variable (eFigure 3).

### Sensitivity analysis

In the first sensitivity analysis, we delayed the beginning of the protocols by one week for the Pfizer-BioNTech vaccine (early: ≤23 days; recommended: 24-32 days; allowable: 33-49 days; late: ≥50 days), making the duration and timing of each protocol exactly the same for both vaccines. Second, we ended the follow-up period at the earliest of SARS-CoV-2 infection, protocol nonadherence, or 180 days after the first dose administration (instead of the end of the study period in the main analysis). This is because we excluded individuals who received their second dose >180 days after their first dose (eMethod 1). Lastly, we excluded 1,335,643 people (21.8%) with missing information on sex, race, ethnicity, and/or public health district, because their infection data were likely missing (eFigure 3).

### Software

All analyses were conducted with R (R Center for Statistical Computing; Vienna, Austria) v4.2.1. The survival analysis was conducted using the ‘survival’ package.^21^

### Ethics statements

This activity was determined by the GDPH Institutional Review Board to be non-research and consistent with public health surveillance as per title 45 code of Federal Regulations 46.102(l)(2).

## RESULTS

### Descriptive statistics

There were 6,128,364 recipients of mRNA COVID-19 vaccines in Georgia who were included in our analysis (Table 1). Of these, 517,966 (8.5%) people had confirmed SARS-CoV-2 infection before vaccination, 26,255 (0.4%) people had infection between the first and second doses, and 388,119 (6.3%) people had infection after the second dose (eTable 2). Of 5,350,766 individuals who had completed the primary series during the study period, 38,539 (0.7%) people received their second dose before the recommended interval, 4,337,660 (81.1%) received within the recommended interval, 834,219 (15.6%) received within the late-but-allowable interval, and 140,348 (2.6%) received within the late interval. Of 777,598 people who had received the first dose but not their second dose by the end of the study period, 717,051 (92.2%) were classified as the late group in the analysis because ≥43 days for Pfizer-BioNTech and ≥50 days for Moderna had passed since their first dose administration. White and Asian individuals were more likely to receive the second dose during the recommended interval compared to other racial groups (eTable 3).

**Table 1.** Characteristics of the vaccine recipients in Georgia, United States, December 2020-March 2022 (N=6,128,364).

| Characteristics | No. (%) or median |
| --- | --- |
| **Sex** |  |
| Female | 3,294,046 (53.8%) |
| Male | 2,772,105 (45.2%) |
| Unknown | 62,213 (1.0%) |
| **Race** |  |
| White | 3,026,177 (49.4%) |
| Black | 1,636,871 (26.7%) |
| Asian | 360,254 (5.9%) |
| AIAN | 22,496 (0.4%) |
| NHPI | 15,020 (0.2%) |
| Other | 850,033 (13.9%) |
| Unknown | 217,513 (3.5%) |
| **Ethnicity** |  |
| Hispanic | 521,415 (8.5%) |
| Non-Hispanic | 5,157,748 (84.2%) |
| Unknown | 449,201 (7.3%) |
| **Age (in years)** | 47.0 |
| **Interval between the 1st and 2nd doses** |  |
| Recommended | 4,337,660 (70.8%) |
| Early | 38,539 (0.6%) |
| Late-but-allowable | 834,219 (13.6%) |
| Late | 140,348 (2.3%) |
| Late (No 2nd dose) | 717,051 (11.7%) |
| Missing | 60,547 (1.0%) |
| **Vaccine manufacturer** |  |
| Moderna | 2,337,570 (38.1%) |
| Pfizer-BioNTech | 3,790,794 (61.9%) |
| **Infection prior to vaccination** |  |
| No | 5,610,398 (91.5%) |
| Yes | 517,966 (8.5%) |
| **Public health district** |  |
| Missing | 956,675 (15.6%) |
| 01-1 | 262,704 (4.3%) |
| 01-2 | 225,202 (3.7%) |
| 02-0 | 334,130 (5.5%) |
| 03-1 | 522,751 (8.5%) |
| 03-2 | 616,773 (10.1%) |
| 03-3 | 133,292 (2.2%) |
| 03-4 | 632,812 (10.3%) |
| 03-5 | 443,544 (7.2%) |
| 04-0 | 389,003 (6.3%) |
| 05-1 | 57,693 (0.9%) |
| 05-2 | 248,382 (4.1%) |
| 06-0 | 222,456 (3.6%) |
| 07-0 | 159,112 (2.6%) |
| 08-1 | 94,846 (1.5%) |
| 08-2 | 166,136 (2.7%) |
| 09-1 | 296,149 (4.8%) |
| 09-2 | 133,407 (2.2%) |
| 10-0 | 233,297 (3.8%) |

| Month and year of 1st dose administration |  |
| --- | --- |
| 2020-12 | 111,878 (1.8%) |
| 2021-01 | 719,154 (11.7%) |
| 2021-02 | 452,909 (7.4%) |
| 2021-03 | 1,330,921 (21.7%) |
| 2021-04 | 918,715 (15.0%) |
| 2021-05 | 433,691 (7.1%) |
| 2021-06 | 237,535 (3.9%) |
| 2021-07 | 297,242 (4.9%) |
| 2021-08 | 518,032 (8.5%) |
| 2021-09 | 298,480 (4.9%) |
| 2021-10 | 149,048 (2.4%) |
| 2021-11 | 224,677 (3.7%) |
| 2021-12 | 204,551 (3.3%) |
| 2022-01 | 152,804 (2.5%) |
| 2022-02 | 60,744 (1.0%) |
| 2022-03 | 17,983 (0.3%) |

Abbreviations: AIAN, American Indian and Alaska Native Resources; NHIS, Native Hawaiian and Pacific Islander.
Intervals between the 1st and 2nd doses: the “early” interval is ≤16 days for Pfizer-BioNTech and ≤23 days for Moderna; the “recommended” interval is 17-25 days for Pfizer-BioNTech and 24-32 days for Moderna; the “late-but-allowable” interval is 26-42 days for Pfizer-BioNTech and 33-49 days for Moderna; the “late” interval is ≥43 days for Pfizer-BioNTech and ≥50 days for Moderna.

### Trial emulation: clone-censor weight analysis

The weighted risk of SARS-CoV-2 infection was similar across all protocols until 25-30 days after the first dose administration (0.57-0.58% on Day 30 since the first dose administration for all protocols) (eFigure 4). Between Day 30 and 60 since the first dose administration, the weighted risk of infection was lowest under the FDA-recommended protocol (RR=1.08 (95% CI: 1.07-1.10) under the late-but-allowable and 1.14 (1.12-1.16) under the late protocol on Day 50 since the first dose administration, compared to the FDA-recommended protocol). After Day 60, the late-but-allowable protocol had the lowest weighted risk of infection (RR=0.83 (95% CI: 0.82-0.84) under the late-but-allowable and 1.10 (1.08-1.12) for the late protocol on Day 120 since the first dose administration, compared to the FDA-recommended protocol). The late protocol consistently had the highest weighted risk of infection.

Next, we ran the analysis separately for Pfizer-BioNTech recipients and Moderna recipients. For Pfizer-BioNTech recipients, the weighted risk of infection was considerably lower under the late-but-allowable protocol (RR=0.79 (95% CI: 0.78-0.81) on Day 120 after the first dose, compared to the FDA-recommended protocol), while the risk under the FDA-recommended protocol was similar to that of the late protocol (Figure 1 and Table 2). For Moderna recipients, the recommended protocol had the lowest risk until about 70 days after the first dose administration, and the late-but-allowable protocol had the lowest risk after that (RR=0.90 (95% CI: 0.87-0.92) on Day 120 after the first dose, compared to the FDA-recommended protocol).

**Table 2.** Inverse probability of censoring-weighted risk of SARS-CoV-2 infection on 50 and 120 days after the first dose administration by protocol, Georgia, United States, 2020-2022.

| Manufacturer | Protocol | 50 days |  | 120 days |  |
| --- | --- | --- | --- | --- | --- |
|  |  | Weighted cumulative risk (95% CI), % | Ratio (95% CI) | Weighted cumulative risk (95% CI), % | Ratio (95% CI) |
| All | Recommended | 0.82 (0.81-0.83) | Ref. | 1.82 (1.80-1.84) | Ref. |
|  | Late-but-allowable | 0.89 (0.88-0.89) | 1.08 (1.07-1.10) | 1.51 (1.49-1.53) | 0.83 (0.82-0.84) |
|  | Late | 0.93 (0.92-0.94) | 1.14 (1.12-1.16) | 2.01 (1.98-2.04) | 1.10 (1.08-1.12) |
| Pfizer-BioNTech | Recommended | 0.93 (0.92-0.95) | Ref. | 2.14 (2.12-2.16) | Ref. |
|  | Late-but-allowable | 1.01 (0.99-1.03) | 1.08 (1.06-1.10) | 1.70 (1.67-1.73) | 0.79 (0.78-0.81) |
|  | Late | 1.07 (1.05-1.09) | 1.14 (1.12-1.16) | 2.21 (2.18-2.25) | 1.03 (1.01-1.05) |
| Moderna | Recommended | 0.62 (0.61-0.63) | Ref. | 1.16 (1.14-1.18) | Ref. |
|  | Late-but-allowable | 0.72 (0.71-0.74) | 1.16 (1.14-1.18) | 1.04 (1.02-1.07) | 0.90 (0.87-0.92) |
|  | Late | 0.73 (0.71-0.76) | 1.17 (1.15-1.21) | 1.44 (1.41-1.55) | 1.24 (1.21-1.33) |

Abbreviations: CI, confidence interval.
Intervals between the 1st and 2nd doses: the “recommended” interval is 17-25 days for Pfizer-BioNTech and 24-32 days for Moderna; the “late-but-allowable” interval is 26-42 days for Pfizer-BioNTech and 33-49 days for Moderna; the “late” interval is $\geq 43$ days for Pfizer-BioNTech and $\geq 50$ days for Moderna.

### Cox PH analysis

Individuals who received their second dose within the recommended interval had the lower hazard of infection after the second dose administration compared to the individuals who received their second dose within the late-but-allowable (aHR=1.047 (95% CI: 1.037-1.057)) and the late (aHR=1.457 (1.423-1.493)) intervals (Table 3). The same pattern was observed after running the models separately for Pfizer-BioNTech and Moderna recipients.

**Table 3.** Estimated hazard ratio (95% confidence interval) for the time from the date of 2nd dose mRNA COVID-19 vaccination to the date of infection or censoring from the **<u>multivariable</u>** Cox proportional hazard model.

| Dosing schedules | Adjusted hazard ratio (95% CI) |  |  |
| --- | --- | --- | --- |
|  | All | Pfizer-BioNTech | Moderna |
| Recommended | Ref. | Ref. | Ref. |
| Early | 1.030 (0.972-1.092) | 1.070 (0.984-1.163) | 0.970 (0.894-1.051) |
| Late-but-allowable | 1.047 (1.037-1.057) | 1.024 (1.012-1.036) | 1.100 (1.080-1.120) |
| Late | 1.457 (1.423-1.493) | 1.381 (1.341-1.422) | 1.585 (1.520-1.652) |

Abbreviations: CI, confidence interval.
Intervals between the 1st and 2nd doses: the “recommended” interval is 17-25 days for Pfizer-BioNTech and 24-32 days for Moderna; the “late-but-allowable” interval is 26-42 days for Pfizer-BioNTech and 33-49 days for Moderna; the “late” interval is $\geq 43$ days for Pfizer-BioNTech and $\geq 50$ days for Moderna.

### Sensitivity analysis

After delaying each interval by one week for the Pfizer-BioNTech vaccine, the weighted risk of infection among Pfizer-BioNTech recipients was estimated to be consistently lowest under the scenario where the second dose was administered from 24-32 days after the first dose (i.e., recommended interval for Moderna) (Figure 2). For the rest of the sensitivity analyses, the results did not meaningfully change for both trial emulation and the Cox PH model (eTable 4-5, eFigure 5-6).

## DISCUSSION

Our novel approach compared the risk of SARS-CoV-2 infection under scenarios where the total study population had followed each of the protocols that varied the timing of second dose administration. The infection risk was compared for the whole course of vaccination, not only after the completion of the primary series but also between the first and second doses. Our findings suggested that mRNA vaccine recipients may gain stronger long-term protection against SARS-CoV-2 infection (as well as disease) by delaying their second dose by approximately one week, especially for Pfizer-BioNTech. However, it is important to note that a delayed second dose leads to a short-term higher risk (during the periods 25-60 days and 30-80 days after the first dose administration for Pfizer-BioNTech and Moderna, respectively, shown in zoomed windows in Figure 1). This short-term higher risk may be particularly relevant if the force of infection in the community is high during that time. We found that a longer delay in the second dose would pose a negative impact in both the short and long-term due to a prolonged time at the lower level of protection induced by a single dose.

Like other more commonly-used methods, the target trial emulation approach handles baseline confounding (by making exact copies of individuals) and informative censoring by protocol non-adherence (with IPCW that account for covariates). However, trial emulation has important advantages. First, our approach yields results with a similar interpretation as those from a trial that randomized people to different vaccine protocols. Second, the method avoids the immortal person-time bias that can occur when the completion of the protocol (i.e., second dose administration) and the start of follow-up (i.e., date of first dose administration) are misaligned. Finally, using this approach, we were able to account for the extended duration of weak-protection time with a single dose under the late protocols, enabling us to evaluate the effectiveness of the whole course of different dosing protocols and providing implications for public health policy. This differs from the more traditional analysis, the Cox PH model, that compared the hazard of infection after the completion of the primary doses across individuals who received the second dose at different timing. Despite its advantages, the trial emulation approach has seldom been used for vaccine evaluation,^14,16^ other than by Butler, *et al*. who used this method to estimate the effects of multi-dose rotavirus vaccine schedules.^18^ We hope that our study serves as another example of the application of the trial emulation approach to vaccine evaluation that would help other researchers to adopt this method.

The trial emulation approach enabled us to understand what would have happened had the total study population followed each of the different dosing protocols, which has policy-relevant implications since a vaccination strategy would be recommended prior to receipt of the first dose. Interestingly, for Pfizer-BioNTech, the risk of infection under the FDA-recommended protocol was similar to that under the late protocol, and the late-but-allowable protocol conferred stronger long-term protection. The sensitivity analysis showed that Pfizer-BioNTech recipients would gain stronger long-term protection when delaying the second dose administration by a week, providing public health implications.

Limitations of our study include that the reported data on vaccination and test results may not be perfectly accurate. We could not analyze outcomes other than SARS-CoV-2 infection, such as death and hospitalization, because the information was frequently missing from surveillance case reports, and the corresponding dates associated with those outcomes were also unreliable. We could not adjust for some important confounding variables, such as comorbidities, employment status, use of non-pharmaceutical interventions (e.g., masking), and results of at-home testing, due to the lack of data. The availability of at-home tests changed over time, especially around late 2021 and early 2022 and the rate of at-home testing may have differed across interdose interval groups, although the difference might be smaller than that between vaccinated and unvaccinated groups.

Our study was able to show how the effectiveness of the mRNA COVID-19 vaccines against SARS-CoV-2 infection varied by the timing of the second dose administration among the general population, providing policy-relevant implications. Delaying the timing of the second dose administration by approximately one week may help to reduce the risk of SARS-CoV-2 infection in the longer term, especially for Pfizer-BioNTech mRNA vaccine. As the COVID-19 pandemic is currently not at its peak, delayed protocols may become even more optimal, as they would not be countered by high vulnerability during the interdose interval. The evaluation for multi-dose vaccination campaigns should be conducted early and periodically to provide evidence of vaccine effectiveness as an outbreak evolves.

## Data Availability

Individual-level data on COVID-19 test results from the Georgia State Electronic Notifiable Disease Surveillance System (SendSS) and COVID-19 vaccination from the Georgia Department of Public Health (GDPH) are also not publicly available, but aggregated data are available on the GDPH COVID-19 website.

## Acknowledgements

The authors thank the Emory COVID-19 Response Collaborative, which is funded by a grant from the Robert W. Woodruff Foundation, for supporting this study and the Georgia Department of Public Health, especially Dr. Laura Edison, Dr. Amanda Jara, and Elizabeth Smith, for sharing the data.

## Author contributions

All authors had full access to all of the data in the study and take responsibility for the integrity of the data and the accuracy of the data analysis.

- Concept and design: E.R.M.
- Data acquisition: P.H.
- Data analysis: K.S.
- Data interpretation: All authors
- Drafting of the manuscript: K.S.
- Critical revision of the manuscript for important intellectual content: All authors
- Statistical analysis: K.S., E.R.M., A.B.
- Administrative, technical, or material support: All authors
- Supervision: E.R.M.

## SUPPLEMENTARY MATERIAL (METHODS)

### eMethods 1: Excluded individuals

We excluded 4,374 (0.1% of mRNA COVID-19 vaccine recipients) people who received their second dose ≤3 days after their first dose because of likely data entry errors. We also excluded 89,885 (1.4%) individuals who received their second dose more than 180 days after their first dose since an interdose interval of that length or longer is unlikely to be recommended and because individuals who received their second dose beyond this time likely received a booster dose at that time while their true second dose was received outside of Georgia or otherwise misrecorded (eFigure 1). Children <5 years of age were excluded as they were not eligible for COVID-19 vaccination during our study period and their primary dosing schedule was different from those for people ≥5 years of age. Recipients of non-mRNA COVID-19 vaccines (e.g., Janssen (Johnson & Johnson) vaccine and Novavax vaccine) were not included in the study.

### eMethods 2: Examples of cloning

In eFigure 2, individual A received the second dose within the recommended interval, and thus, it was followed up until the end of the study period in the copy for the “recommended” protocol (i.e., survival time T days), while it was censored on the day of the second dose administration (Day 21) in the copies for the “late but allowable” and “late” protocols (i.e., survival time 21 days). Individual B was censored on the date of second dose administration (Day 13) in all copies. Individual C was censored on the last day of the recommended interval (Day 25) in the recommended protocol copy (i.e., survival time 25 days), while it was followed up until the day of COVID-19 infection in the “late but allowable” protocol copy (i.e., survival time 36 days) and it was censored on the day of second dose administration (Day 31) in the copy for the “late” protocol (i.e., survival time 31 days). Individual D received the second dose during the late interval, and thus, it was censored on the last day of the recommended interval in the recommended protocol copy (i.e., survival time 25 days) and on the last day of the late but allowable interval in the “late but allowable” protocol copy (i.e., survival time 42 days), while it was followed up until the end of the study period in the “late” protocol copy (i.e., survival time T days). Individual E was followed up until the day of COVID-19 infection (Day 7) in all copies (i.e., survival time 7 days).

## SUPPLEMENTARY TABLES

**eTable 1.**
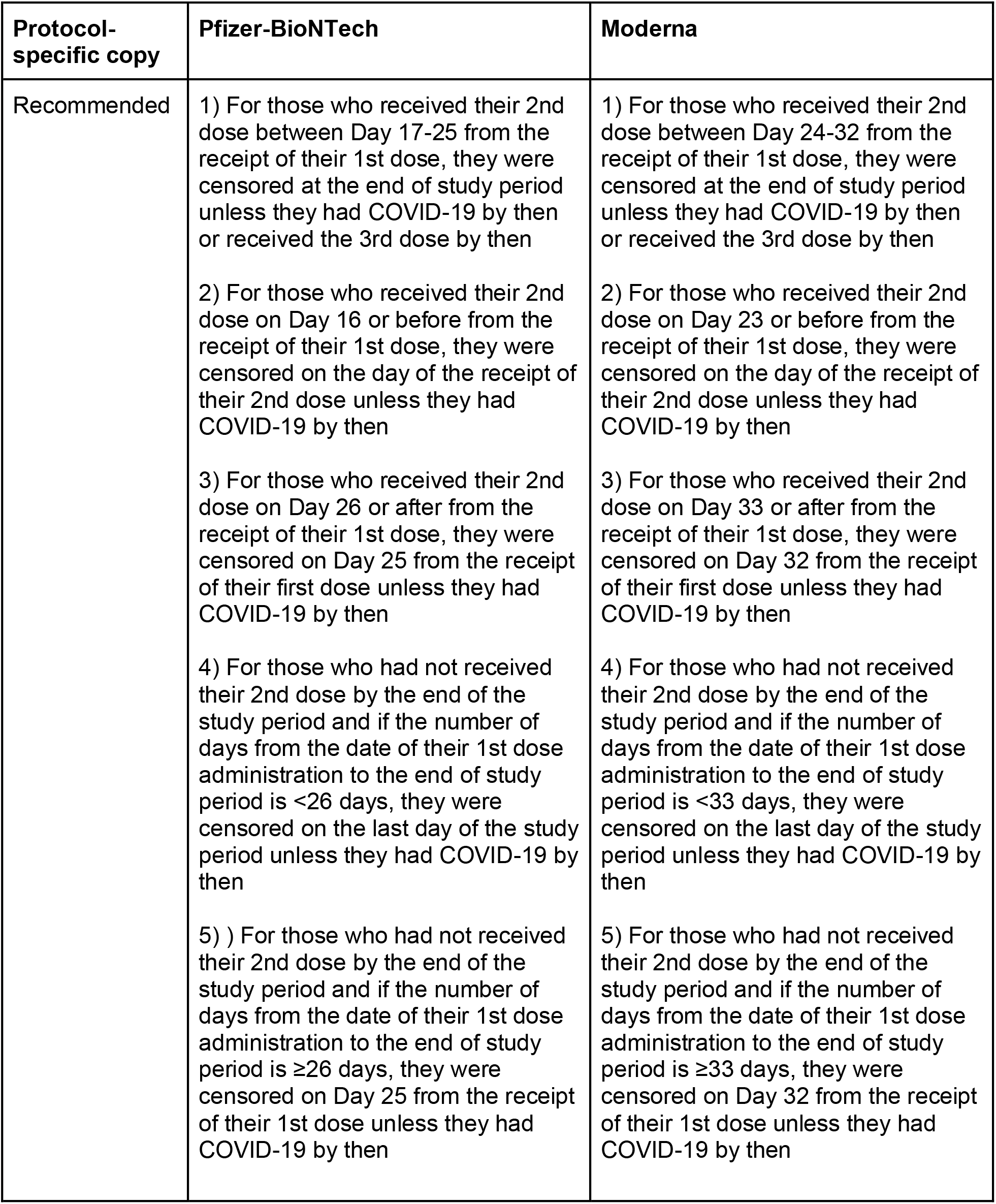
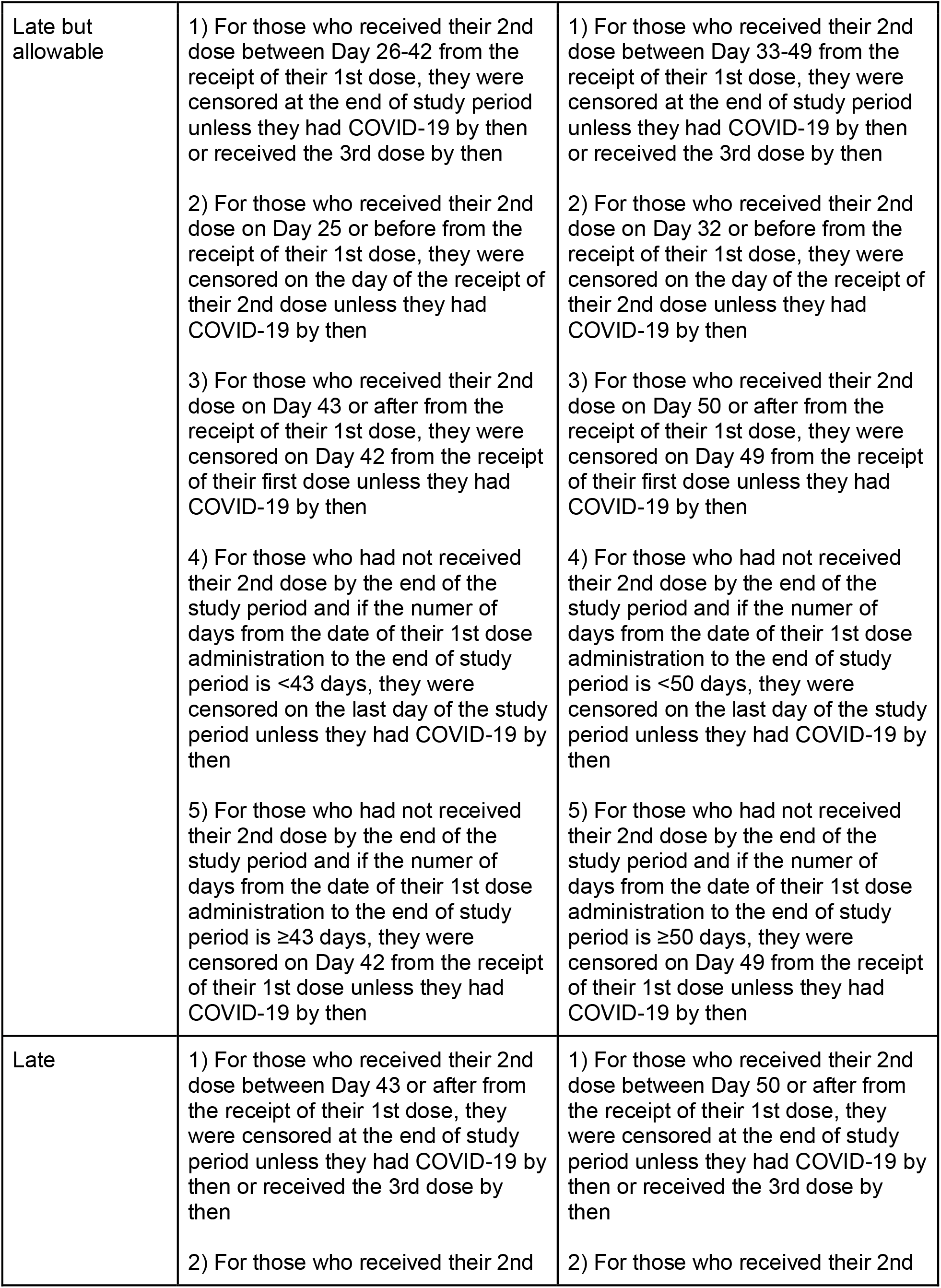
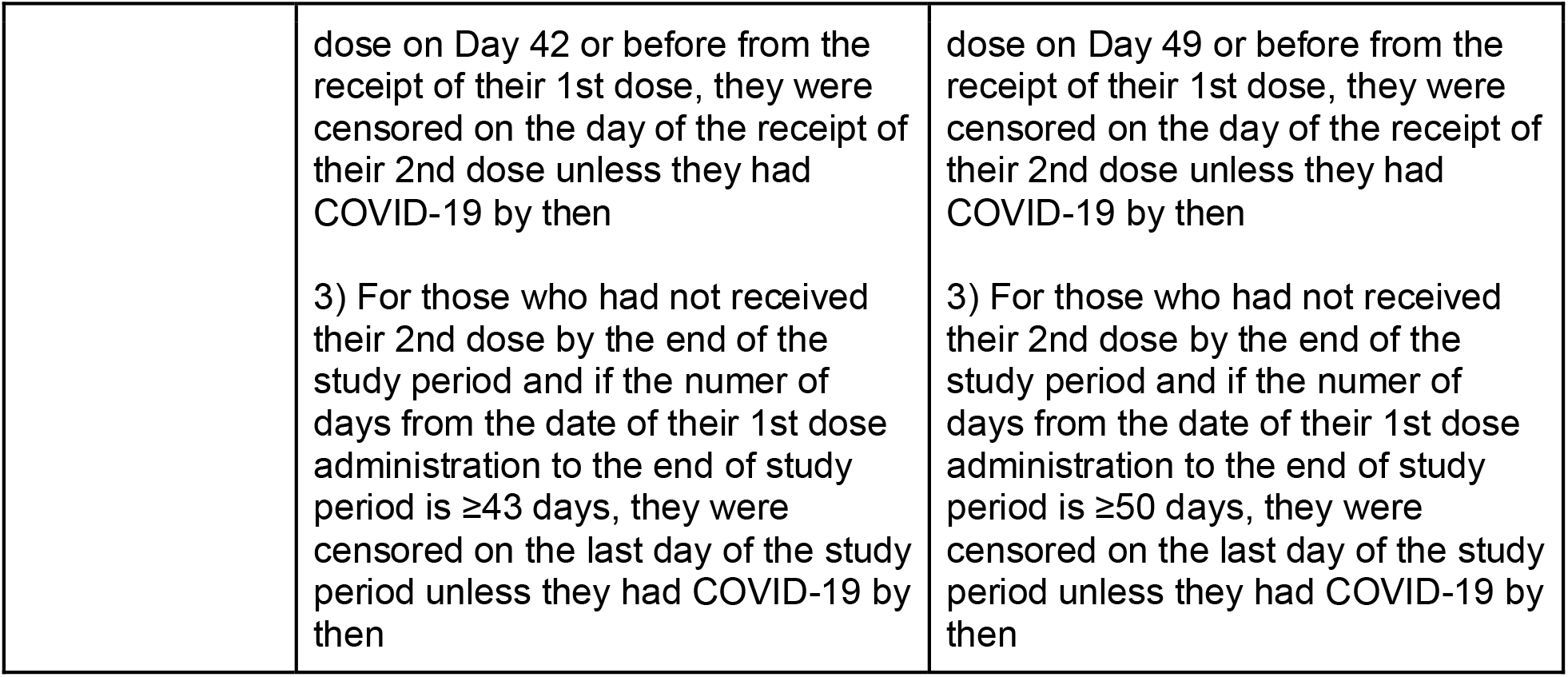
Condition on censoring in each of three copies of the longitudinal dataset corresponding to the mRNA COVID-19 vaccine protocols

**eTable 2.**
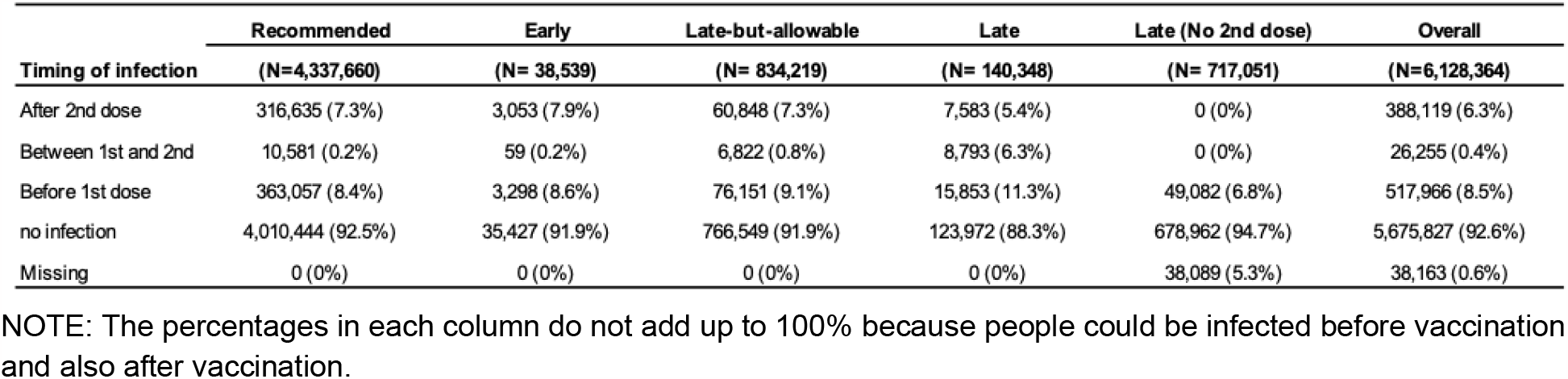
Timing of SARS-CoV-2 infection relative to vaccination by intervals between the first and second doses of mRNA COVID-19 vaccines.

**eTable 3.**
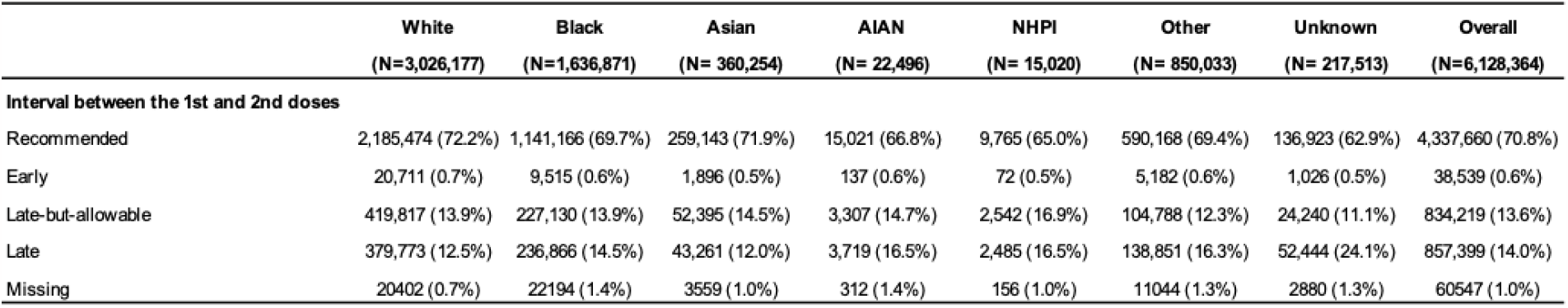
mRNA COVID-19 vaccines interdose intervals by race.

**eTable 4.**
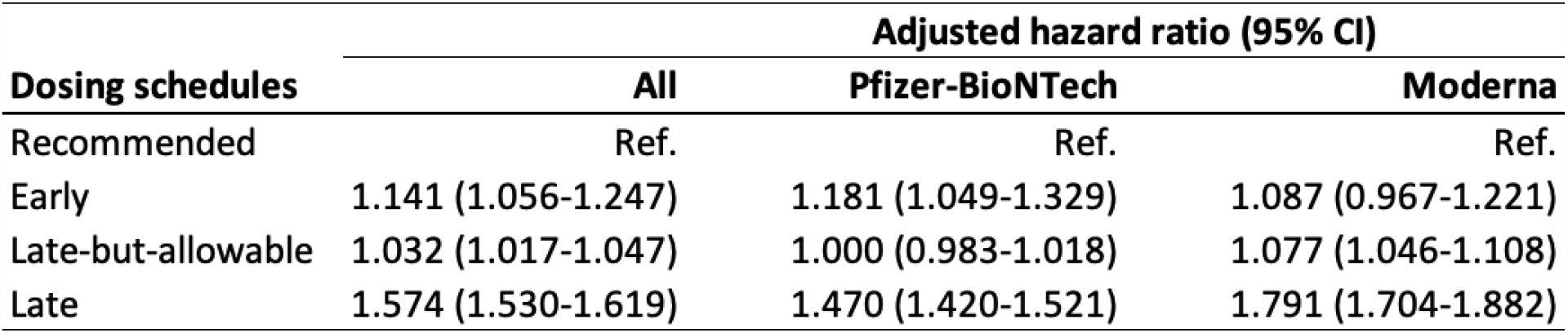
Results of the second sensitivity analysis (ending the follow-up period at the earliest of SARS-CoV-2 infection, protocol nonadherence, or 180 days after the first dose administration): Estimated hazard ratio (95% confidence interval) for the time from the date of 2nd dose mRNA COVID-19 vaccination to the date of infection or censoring from the multivariable Cox proportional hazard model.

**eTable 5.**
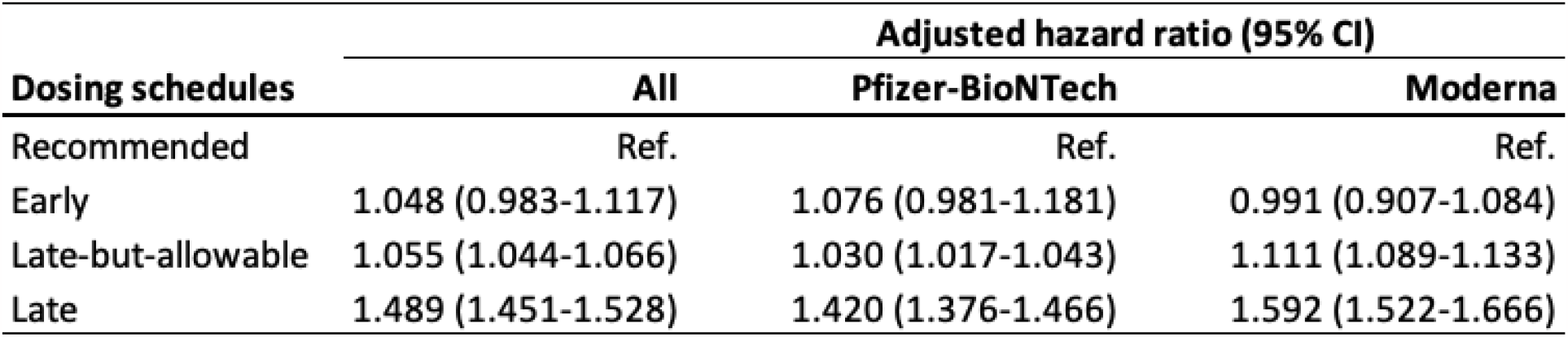
Results of the third sensitivity analysis (excluding individuals with unknown characteristics): Estimated hazard ratio (95% confidence interval) for the time from the date of 2nd dose mRNA COVID-19 vaccination to the date of infection or censoring from the multivariable Cox proportional hazard model.

## SUPPLEMENTARY FIGURES

**eFigure 1.**
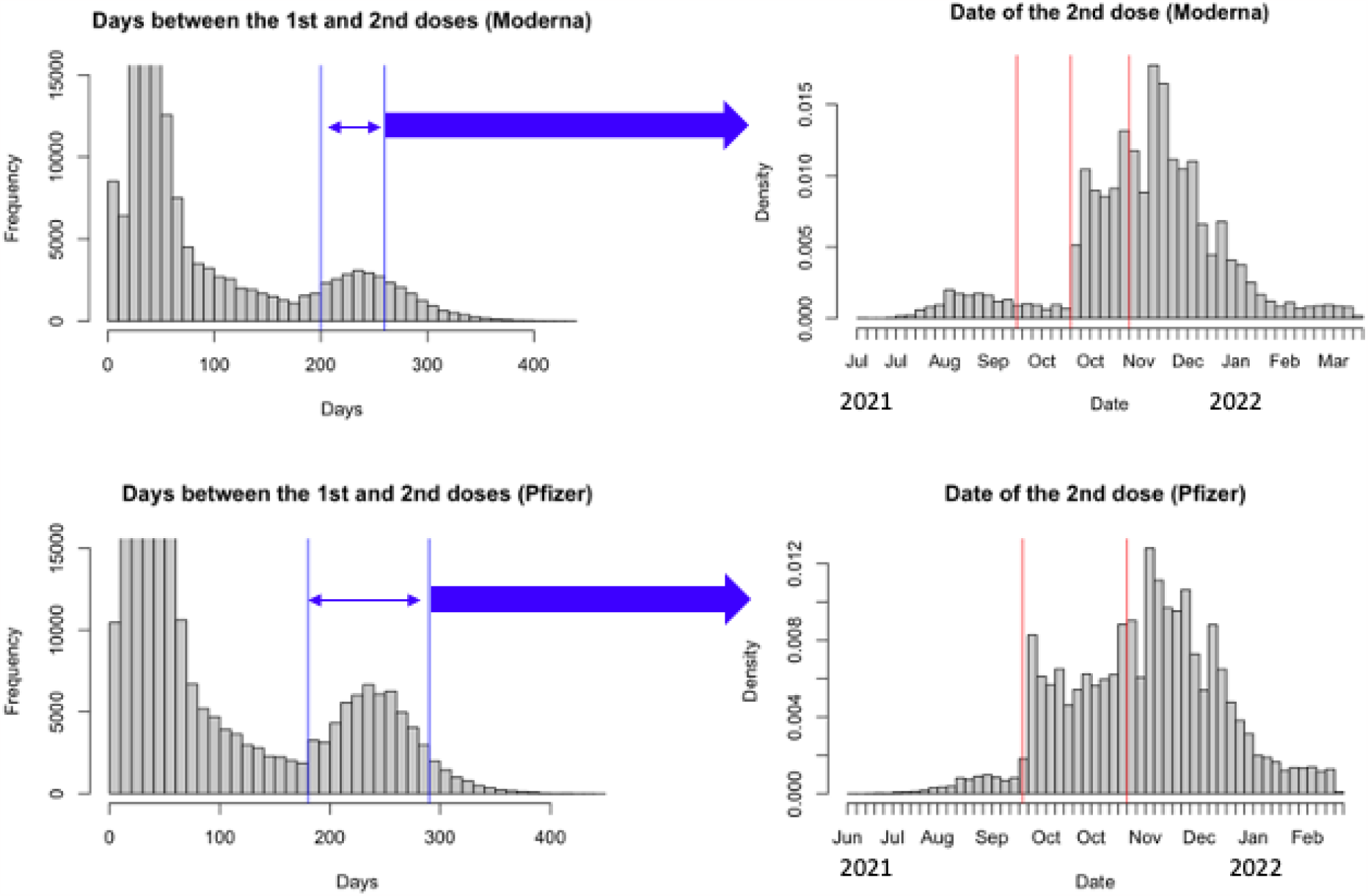
Date of the second dose administration for people who received the 2nd dose >180 days after their 1st dose administration. Red vertical lines: 2021-09-22: PFR booster available for high-risk groups 2021-10-20: MOD booster available for high-risk groups 2021-11-19: Booster available for general public

**eFigure 2.**
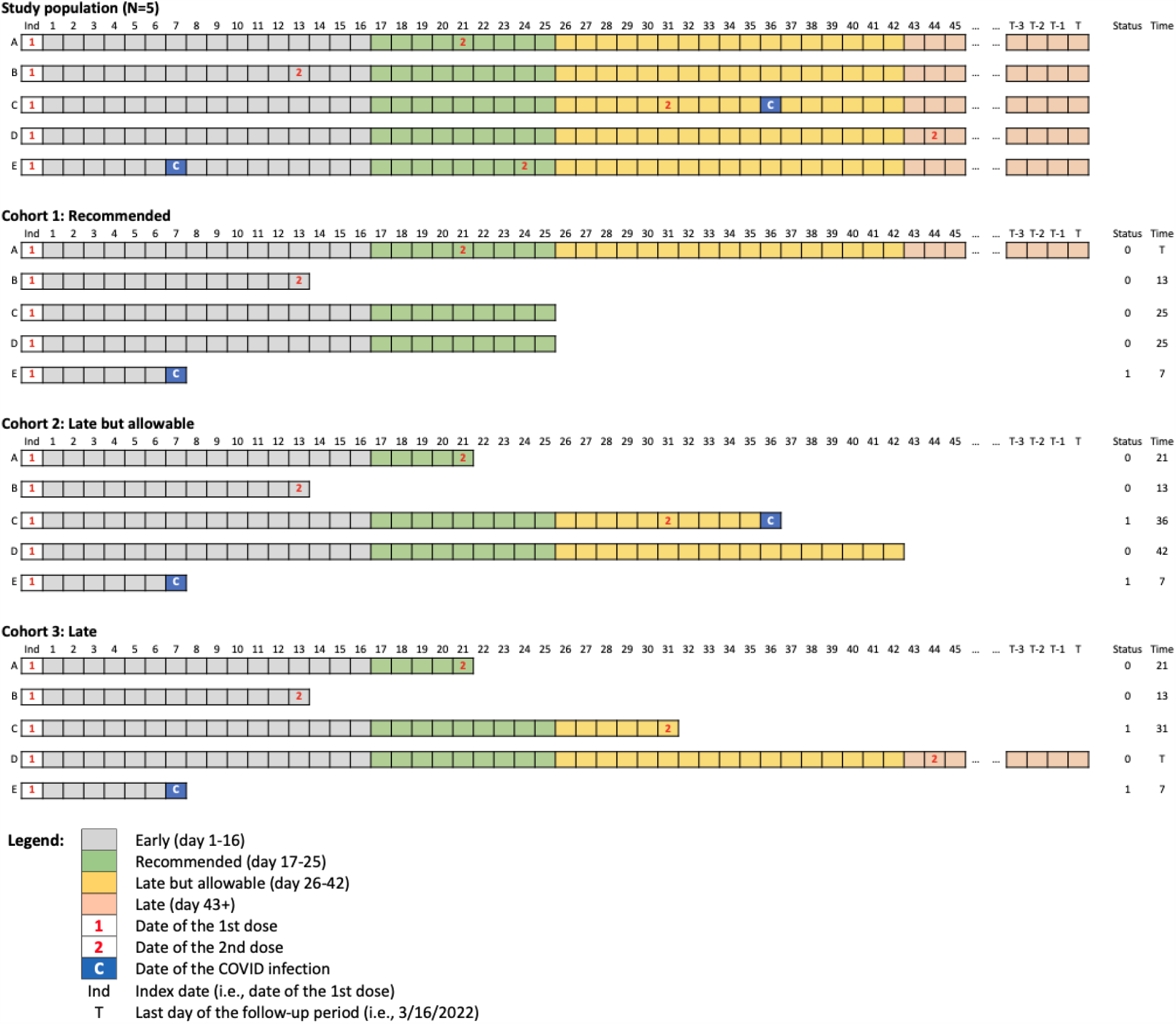
Example of the longitudinal dataset for five vaccine recipients and its three copies corresponding to different vaccine protocols (recommended, late but allowable, and late) for Pfizer-BioNTech mRNA COVID-19 vaccine.

**eFigure 3.**
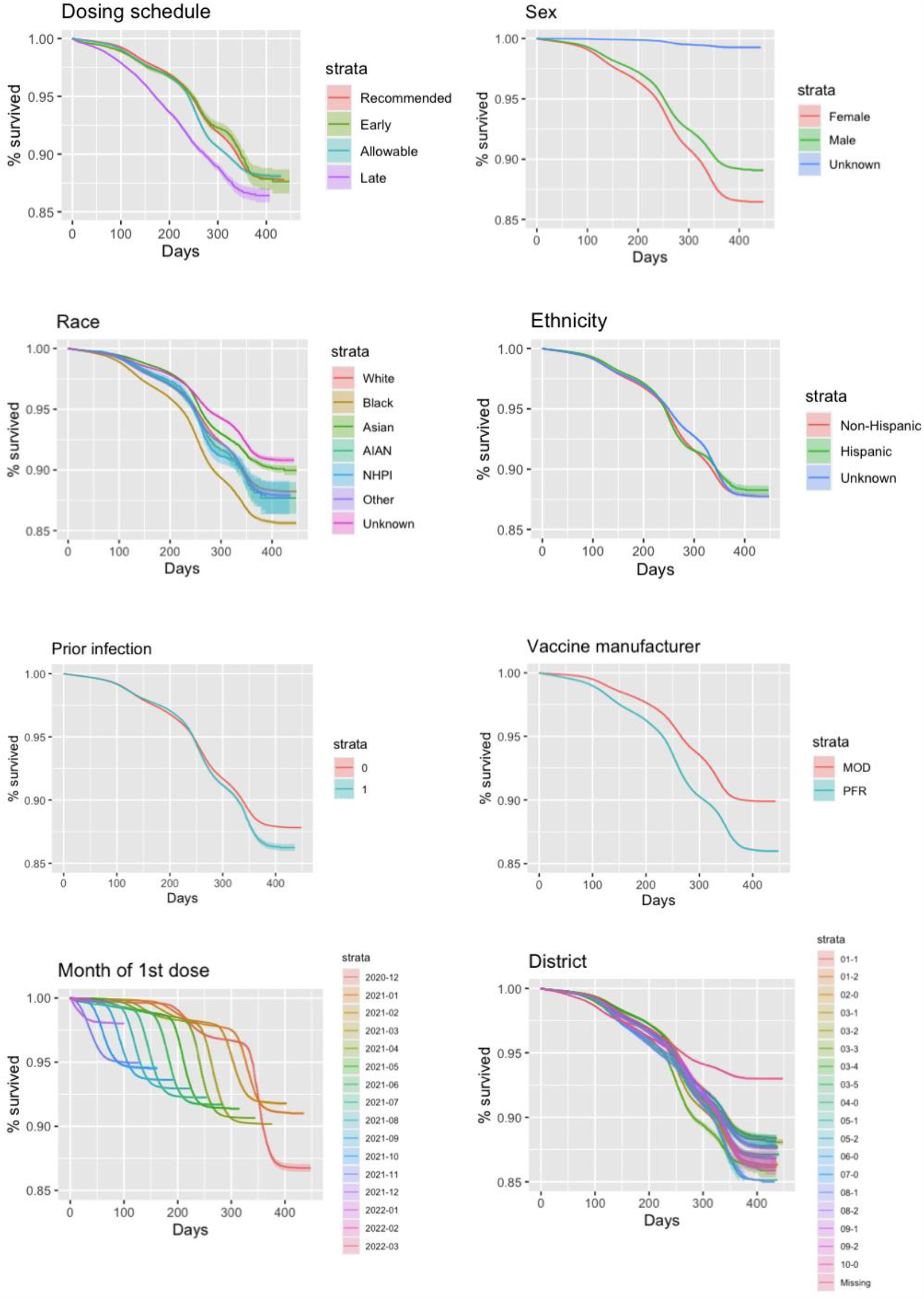
Kaplan-Meier survival curves comparing the time from the second dose administration to SARS-CoV-2 infection by the exposure variable and covariates. Tic marks for censoring are omitted from the figures.

**eFigure 4.**
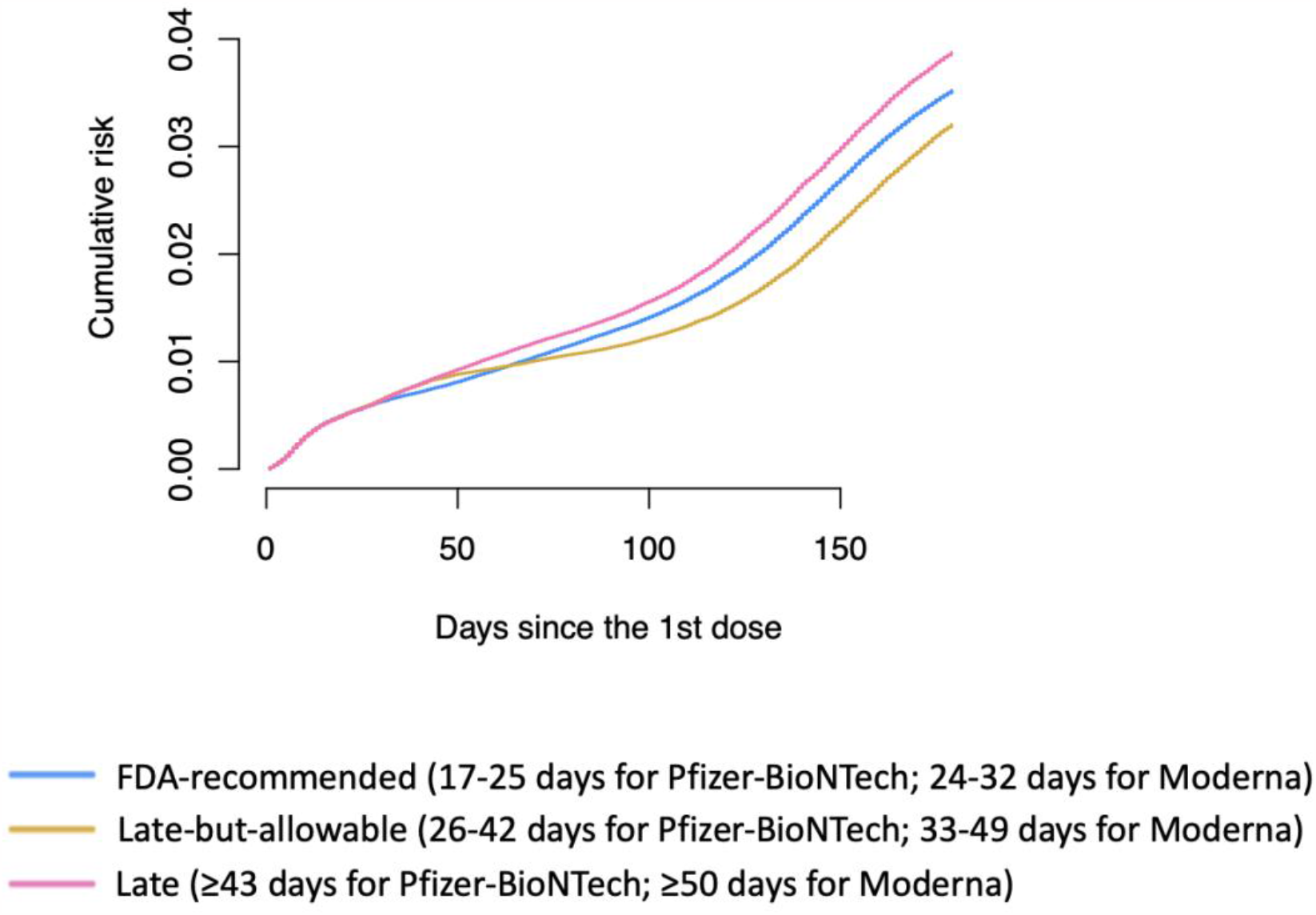
Estimates of inverse probability of censoring-weighted cumulative risk functions of SARS-CoV-2 infection by protocol.

**eFigure 5.**
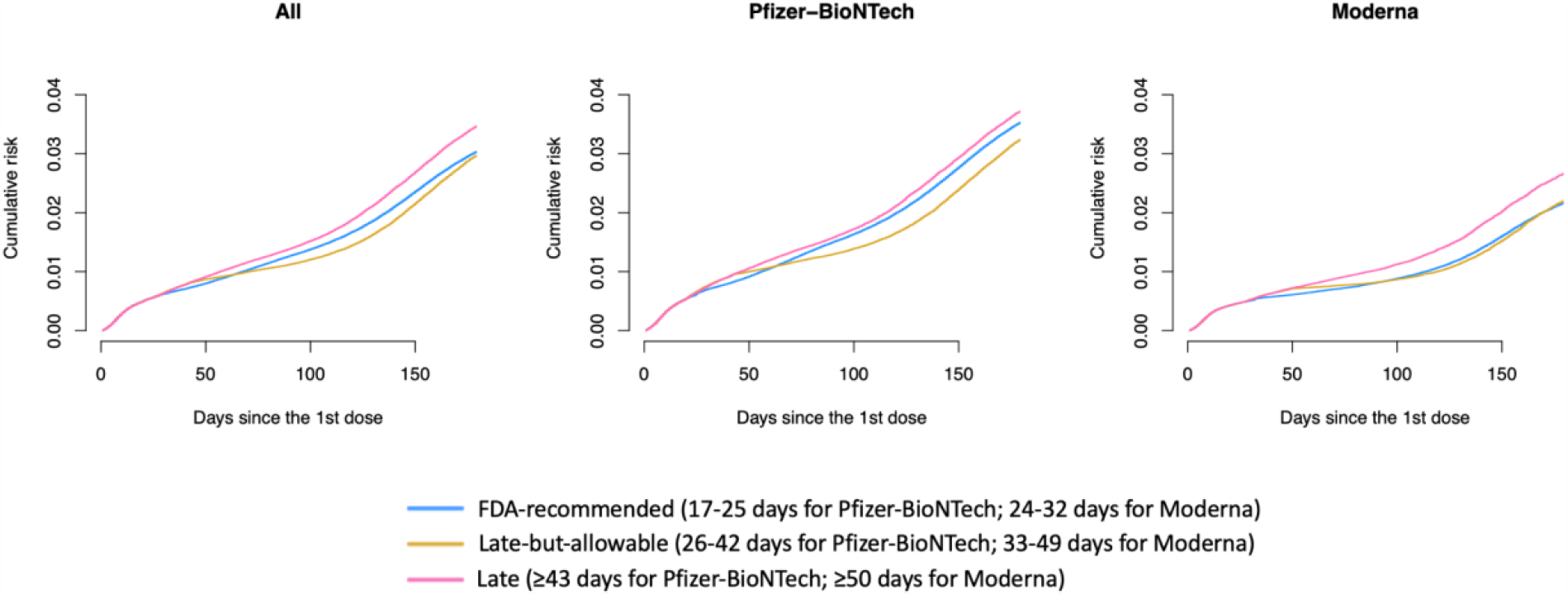
Results of the second sensitivity analysis (ending the follow-up period at the earliest of SARS-CoV-2 infection, protocol nonadherence, or 180 days after the first dose administration): Estimates of inverse probability of censoring-weighted cumulative risk functions of SARS-CoV-2 infection by protocol.

**eFigure 6.**
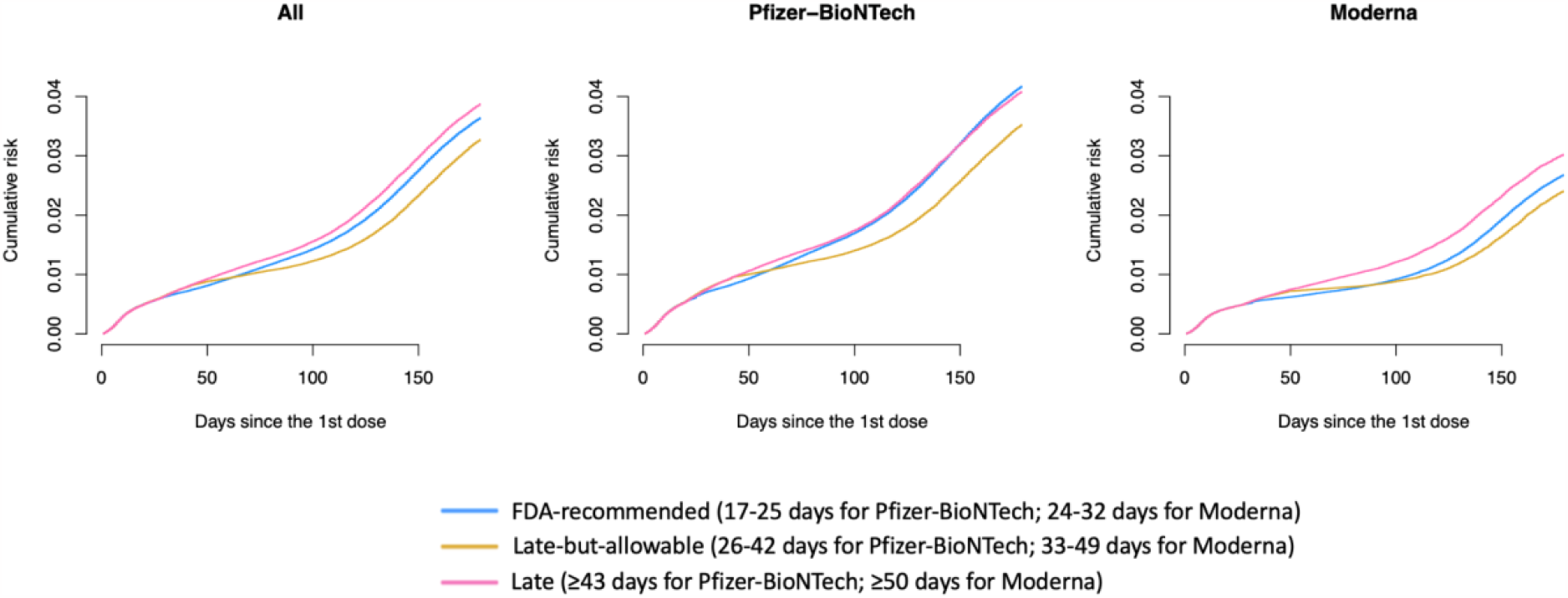
Results of the third sensitivity analysis (excluding individuals with unknown characteristics): Estimates of inverse probability of censoring-weighted cumulative risk functions of SARS-CoV-2 infection by protocol.

